# A multiplex microsphere IgG assay for SARS-CoV-2 using ACE2-mediated inhibition as a surrogate for neutralization

**DOI:** 10.1101/2020.10.05.20203976

**Authors:** Andrew Cameron, Claire A. Porterfield, Larry D. Byron, Jiong Wang, Zachary Pearson, Jessica L. Bohrhunter, Anthony B. Cardillo, Lindsay Ryan-Muntz, Ryan A. Sorensen, Mary T. Caserta, Steven Angeloni, Dwight J. Hardy, Martin S. Zand, Nicole D. Pecora

## Abstract

The COVID-19 pandemic has highlighted the challenges inherent to the serological detection of a novel pathogen such as SARS-CoV-2. Serological tests can be used diagnostically and for surveillance, but their usefulness depends on their throughput, sensitivity and specificity. Here, we describe a multiplex fluorescent microsphere-based assay, 3Flex, that can detect antibodies to three major SARS-CoV-2 antigens—spike (S) protein, the spike ACE2 receptor-binding domain (RBD), and nucleocapsid (NP). Specificity was assessed using 213 pre-pandemic samples. Sensitivity was measured and compared to the Abbott™ ARCHITECT™ SARS-CoV-2 IgG assay using serum samples from 125 unique patients equally binned (*n* = 25) into 5 time intervals (≤5, 6 to 10, 11 to 15, 16 to 20, and ≥21 days from symptom onset). With samples obtained at ≤5 days from symptom onset, the 3Flex assay was more sensitive (48.0% *vs*. 32.0%), but the two assays performed comparably using serum obtained ≥21 days from symptom onset. A larger collection (*n* = 534) of discarded sera was profiled from patients (*n* = 140) whose COVID-19 course was characterized through chart review. This revealed the relative rise, peak (S, 23.8; RBD, 23.6; NP, 16.7; in days from symptom onset), and decline of the antibody response. Considerable interperson variation was observed with a subset of extensively sampled ICU patients. Using soluble ACE2, inhibition of antibody binding was demonstrated for S and RBD, and not for NP. Taken together, this study described the performance of an assay built on a flexible and high-throughput serological platform that proved adaptable to the emergence of a novel infectious agent.

## INTRODUCTION

The β-coronavirus SARS-CoV-2 is the causative agent of the coronavirus disease 2019 (COVID-19) pandemic. As SARS-CoV-2 continues to move through the global population, serological testing is vitally important to understand both the degree of spread (seroprevalence) and the characteristics of the immune response in terms of antibody kinetics, dominant antigens, and immunological significance.

A number of antibody tests have been granted emergency use authorization (EUA) by the FDA ^1^. These include rapid lateral flow devices (LFAs), enzyme-linked immunoassays (ELISAs), chemiluminescent immunoassays (CMIAs), and fluorescence microsphere immunoassays (FMIA). The majority of these tests are qualitative and typically target either the SARS-CoV-2 spike (S) or nucleocapsid (NP) proteins. Likewise, these same targets were of interest in the development of serological assays for SARS-CoV-1 ^2-6^.

The S protein is a ∼180 kDa glycosylated homotrimer that extends from the viral surface and initiates host cell entry ^7^. The extracellular region of S is organized into S1 and S2, with S1 comprising the outermost region and containing the receptor binding domain (RBD) for the human ACE2 receptor ^8-11^. Anti-S antibodies can neutralize SARS-CoV-2 in cell culture ^7,8,12-15^, as can anti-S antibodies against SARS-CoV-1 ^3^. Thus, S and RBD are of great interest as targets for both immunoassays and vaccines due to their interactions with the human host ^16^. As a component of serological assays, S is incorporated as either the full extracellular domain (ECD), the outermost S1 domain, or the receptor-binding domain (RBD). Additionally, the abundance and antigenicity of NP have made it an attractive target for both diagnostic and vaccine work ^17-19^. NP has roles in viral replication, transcription, and packaging of the viral genome ^20,21^.

Beyond exposure, questions surrounding the serological response to SARS-CoV-2 are becoming more nuanced, and include long-term antibody kinetics, measuring anamnestic responses to reinfection or vaccination, and assessment of plasma donors. For these, both qualitative and quantitative/semi-quantitative serological assays that target multiple antigens and/or have the ability to assess the neutralization potential of SARS-CoV-2 antibodies are of great interest. To date, only a handful of assays with FDA EUA incorporate both the NP and S antigens, including technologies such as LFAs (e.g. Assure COVID-19 IgG/IgM Rapid Test Device, CareStart COVID-19 IgM/IgG, Cellex qSARS-CoV-2 IgG/IgM Rapid Test), CLIAs (e.g. Diazyme DZ-Lite SARS-CoV-2 IgM/IgG, Vibrant COVID-19 Ab Assay), and FMIAs (Luminex Inc. xMAP SARS-CoV-2 Multi-Antigen IgG Assay) ^1^. None of the current EUA assays incorporate an assessment of antibody-mediated neutralization of SARS-CoV-2 S protein binding to the ACE2 receptor.

In the current study, we utilized a laboratory-developed FMIA designated SARS-CoV-2 IgG 3Flex (3Flex) built on the Luminex FLEXMAP 3D^®^ system for the simultaneous detection of antibodies to the N, S, and RBD antigens. We assessed its performance using 534 serum samples from 140 SARS-CoV-2-infected individuals from both inpatient and outpatient settings. In addition, we followed the serological response in a subset of individuals over time, and demonstrated longitudinal patterns of antibody levels to each antigen alongside an assessment of neutralization at different time points over the course of disease.

## MATERIALS AND METHODS

### Clinical laboratory setting

The UR Medicine Labs clinical microbiology laboratory in Rochester, NY provides diagnostic services for institutionalized and ambulatory patients (serving several area hospitals, practices, nursing homes, and urgent care providers). During the ongoing COVID-19 global pandemic, diagnostic analyses for SARS-CoV-2 were provided for an expanded patient population in Western NY. This study was approved by the University of Rochester Institutional Review Board (RSRB STUDY00004836).

### Sample collection

Residual serum specimens (*n* = 534) were collected at convenience in April 2020 from 140 unique patients (including outpatients, nursing home residents, and inpatients). All serum specimens were obtained from symptomatic patients positive for SARS-CoV-2 nucleic acid tested with RT-PCR-based assays (cobas^®^ SARS-CoV-2 Test, Simplexa™ COVID-19 Direct, and Xpert^®^ Xpress SARS-CoV-2). Serum was held at 4°C until discard (7 days) or −80°C for long-term storage. Additionally, pre-COVID-19 sera (*n* = 125) from 2019 were used for specificity testing and validation, and included specimens positive for bacterial and viral infections (lyme, syphilis, cytomegalovirus [CMV], Epstein-Barr virus [EBV], and other respiratory illnesses) and autoimmune markers (antinuclear antigen [ANA], and rheumatoid factor).

### Chart review

Patient charts were reviewed by at least two people from a four-person team, and discrepancies were resolved by a third person. Information collected included patient demographics (sex, age), clinical course (including the estimated date of symptom onset, days of hospitalization, ICU admission, death), reported symptoms (including fever, cough, shortness of breath), and comorbidities (including history of smoking, coronary artery disease, chronic obstructive pulmonary disease [COPD], and diabetes). Given the need to assess for the serological response within a known timeframe (days from symptom onset), all patients in this study had symptomatic manifestations of COVID-19. Thus, days from symptom onset was calculated using the collection date for each serum sample. Where possible, RT-PCR cycle threshold (C_t_) values from multiple RT-PCR tests were obtained and assessed alongside the serological response for the patients included in this study.

### Multiplex SARS-CoV-2 IgG microsphere immunoassay development

#### Antigens

The immunoassay was constructed using full-length SARS-CoV-2 NP protein (Cat. Z03480-100), the extracellular domain (ECD) of the S protein (Cat. Z03481-100), and RBD (Cat. Z03483-100), all purchased from GenScript Biotech Corporation, Piscataway, NJ. The S protein (135 kDa; His tag & FLAG^®^ tag) was expressed in *Sf9* insect cells, NP protein (46 kDa; His tag) was expressed in *E. coli*, and RBD (30 kDa; His Tag) was obtained from expression in human cells. The SARS-CoV-2 spike ECD and RBD are both reported to ‘bind with human ACE2 in functional ELISA assay’ by the manufacturer. In addition to the viral antigens, two additional control beads were included: an internal control bead on region 45 (IC45), and a bead coupled with human IgG, as described in the next section.

#### Microsphere-antigen coupling

Protein coupling to magnetic microsphere beads was performed according to the manufacturer’s recommendations using the xMAP^®^ Antibody Coupling (AbC) kit (Luminex, Austin, TX). The S protein was coupled at 10 pM, while NP and RBD were coupled at 100 pM (i.e. equimolar). Beads were also coated with purified human IgG at 0.1 μg/ml (Cat. I2511, Sigma-Aldrich, Inc.). Coupled beads corresponded to bead regions 18 (S), 33 (RBD), 73 (NP), 45 (internal control; IC45), and 66 (human IgG). IgG coupling confirmation was performed using serial dilutions of biotin-SP-conjugated Fcγ fragment-specific goat α-human IgG (Cat. 109-065-098, Jackson ImmunoResearch Laboratories, Inc.) and streptavidin-R-phyocerythrin (PE) conjugate (SAPE, Cat. S21388, Thermo Fisher Scientific). SARS-CoV-2 antigen coupling was confirmed serial dilutions of rabbit-derived antibodies directed against S (Cat. NB100-56047) and NP (Cat. NB100-56049; Novus Biologicals, LLC, Centennial, CO) and detection with PE-labelled goat α-rabbit IgG (H+L) cross-adsorbed secondary antibody (Cat. P2771MP, Thermo Fisher Scientific).

#### Assay procedure

Serum was diluted to 1:2000 in PBS-TBN (PBS: 0.02%; Tween: 0.1%; BSA: 1%; Azide: 0.05%). The assay was performed by incubating a 5-plex bead mix delivering 2,500 beads for each bead region (IC45, each CoV antigen bead, and the IgG-coupled bead) in a 50 µl volume of PBS-TBN with 50 µl of the diluted patient serum for 15 min at 37°C. Beads were washed twice in 150 µl of PBS-TBN with shaking for 2 minutes at 37°C using magnetic separation. Washed beads were incubated with a mixture of 1:4500 biotin-SP-conjugated Fcγ fragment-specific goat α-human IgG and 1 μg/ml SAPE for 15 min at 37°C. Beads were again washed twice in 150 µl of PBS-TBN with shaking for 2 minutes at 37°C and separated magnetically. Beads were re-suspended in 100 µl of PBS-TBN and analyzed on a FLEXMAP 3D^®^ instrument (Luminex, Austin, TX), which provides a measurement of the median fluorescence intensity (MFI) for each analyte on each bead region. For each 96-well plate, patient sera was tested alongside three PBS-TBN blank wells, an IgG-stripped sera control (Cat. IIGGDS100ML, Innovative Research, Inc., Novi, MI), and two sera from patients positive for SARS-CoV-2 by RT-PCR known to be of ‘low’ and ‘high’ positivity with respect to SARS-CoV-2 IgG serology (as determined by the Abbott™ SARS-CoV-2 IgG assay).

### Comparator SARS-CoV-2 IgG testing

Sera used in the validation of the 3Flex assay were tested in parallel according to the package insert for the FDA Emergency Use Authorization (EUA) SARS-CoV-2 IgG assay on the Abbott™ ARCHITECT™ *i*2000SR platform. This assay detects the serological response to SARS-CoV-2 NP.

### Study design and analysis

#### *Multiplex* SARS-CoV-2 IgG *microsphere immunoassay validation*

To establish the required specificity of the assay, and for interference testing, MFI cut-off values were determined by testing a total of 125 ‘pre-COVID’ sera. These comprised sera positive for ANA (*n* = 20), rheumatoid factor (*n* = 10), lyme (*n* = 10), syphilis (*n* = 10), CMV (*n* = 10), EBV (*n* = 10), and non-SARS-CoV-2 respiratory illnesses (*n* = 55). To examine assay performance, sera (*n* = 125) from unique patients (i.e. *n* = 125) positive for SARS-CoV-2 nucleic acid and with a known date of symptom onset were tested. These samples were randomly selected and binned into 5 time intervals corresponding to ≤5, 6 to 10, 11 to 15, 16 to 20, and ≥21 days from symptom onset. Results were interpreted qualitatively and concordance was established with respect to the Abbott™ SARS-CoV-2 IgG assay.

#### *Serological profiling of* SARS-CoV-2 *infection*

MFI values for each antigen from all tests (*n* = 534) were assessed with respect to days from symptom onset. MFI values and corresponding C_t_ values were plotted in GraphPad Prism 8 with a smoothed curve (GraphPad Software, San Diego, CA). Peak values for IgG responses were determined by area under the curve (AUC) analyses. Because some patients were highly represented in the larger data set, a sub-set of randomly selected serum specimens was used for MFI comparisons by time, and for comparisons between select patient populations (i.e. between ICU-admitted and other patients). Where possible, this sub-set included no more than 1 serum specimen assayed from a unique patient for each of the 5 time intervals. This allowed 231 serological tests from 140 unique patients to be examined. Additionally, 9 extensively sampled patients were tested to explore interperson (i.e. between patient) IgG responses and the precision of the assay with repeated measures.

#### ACE2 inhibition

As a proxy for the detection of ‘neutralizing’ titers of antibodies to SARS-CoV-2, the 5-plex bead mix was incubated with soluble recombinant human angiotensin-converting enzyme 2 (ACE2) (Cat. 0192-30, Adipogen Corporation, San Diego, CA) for 2 minutes at 37°C with shaking prior to the addition of sera. A doubling dilution series of ACE2 was used to optimize the concentration needed to produce a ∼50% loss of MFI value for sera tested with and without ACE2. A concentration of 2 μg/ml was selected because MFI values for the spike RBD were reduced by 50% in the majority of SARS-CoV-2 positive samples tested. ACE2 inhibition was also performed for the 9 extensively characterized patients, but only 1 randomly-selected serum sample from each of the 5 time intervals was tested if possible. Inhibition values—given as the residual MFI + ACE2 (%)—are calculated as the percentage of the MFI value in the presence of ACE2 over the MFI value without ACE2.

### Statistical and graphical analysis

Statistical calculations and plotting were performed in Prism 8 (GraphPad Software, San Diego, CA). Fisher’s exact test was used for patient population comparisons. Unless otherwise indicated, error bars indicate mean ± standard deviation.

## RESULTS

### Multiplex SARS-CoV-2 IgG microsphere immunoassay validation

#### Specificity

Specificity was assessed using a set of 213 pre-COVID-19 sera. Serum samples were submitted to the diagnostic laboratory between 10/1/2019 and 2/1/2020 and included those that were negative for all analytes tested, as well as samples that were positive for syphilis, CMV, EBV/Mono, rheumatoid factor, and Lyme disease **(Table 1)**. This also included 55 samples taken from patients within 60 days of an acute respiratory infection. All pre-COVID-19 sera were designated as ‘negative’ and formed the basis of cut-off values to establish positive/negative thresholds used to interpret subsequent testing. The MFI cut-off values for each antigen exceeded 6 standard deviations of the mean of all negative samples included in this specificity assessment.

**Table 1:** 3Flex MFI^*a*^ values of negative pre-COVID-19 serum and serum positive for potential interfering etiologies.

| Sample group | *n* | Spike MFI Average<br>(95% CI) | RBD MFI Average<br>(95% CI) | Nucleocapsid MFI Average<br>(95% CI) |
| --- | --- | --- | --- | --- |
| Pre-COVID-19 | 93 | 140<br>(127-153) | 91<br>(88-93) | 224<br>(159-288) |
| Lyme | 10 | 148<br>(138-157) | 97<br>(92-102) | 564<br>(58-1,069) |
| Syphilis | 10 | 165<br>(148-182) | 105<br>(98-113) | 214<br>(3-284) |
| CMV | 10 | 180<br>(113-247) | 96<br>(91-102) | 230<br>(99-361) |
| EBV/Mono | 10 | 187<br>(134-241) | 109<br>(102-117) | 265<br>(165-365) |
| Acute viral respiratory illness | 55 | 136<br>(121-151) | 91<br>(90-93) | 165<br>(107-223) |
| ANA | 20 | 178<br>(153-202) | 101<br>(97-104) | 348<br>(162-535) |
| Rheumatoid Factor | 10 | 149<br>(134-164) | 99<br>(94-104) | 476<br>(80-873) |

<sup>a</sup> MFI: median fluorescent intensity.
Abbreviations: CMV, cytomegalovirus; EBV/Mono, Epstein-Barr virus mononucleosis; ANA, anti-nuclear antigen.

#### Sensitivity

Serum samples from patients with confirmed COVID-19 were categorized by days from symptom onset (≤5, 6 to 10, 11 to 15, 16 to 20, and ≥21 days; *n* = 25 each) and evaluated with both the 3Flex assay and the Abbott™ ARCHITECT™ SARS-CoV-2 IgG assay (EUA) **(Table 2)**. If any of the 3Flex markers (S, RBD, NP) was positive then the overall interpretation of the assay was positive. With serum obtained ≤5 days from symptom onset, the 3Flex assay demonstrated a 48.0% (12/25) positivity rate, compared to 32.0% for the ARCHITECT™ SARS-CoV-2 IgG assay (8/25) **(Table 2)**. In 3/4 of the discrepant samples, the 3Flex assay was positive due to the S target alone, while in 1/4 all three targets were positive **(Table S1)**. By 16 to 20 days from symptom onset the two assays showed comparable positivity rates (92%) which were also observed for samples obtained ≥21 days from symptom onset. Although the 3Flex assay is reported with a positive or negative qualitative result, MFI values increased from an average of 2,305.88 (S), 1,168.40 (RBD), and 3,380.28 (NP) in the ≤5 days from symptom onset category to 24,859.64 (S), 14,963.76 (RBD), and 20,801.24 (NP) in the ≥21 days from symptom onset category **(Table 2)**.

**Table 2:**
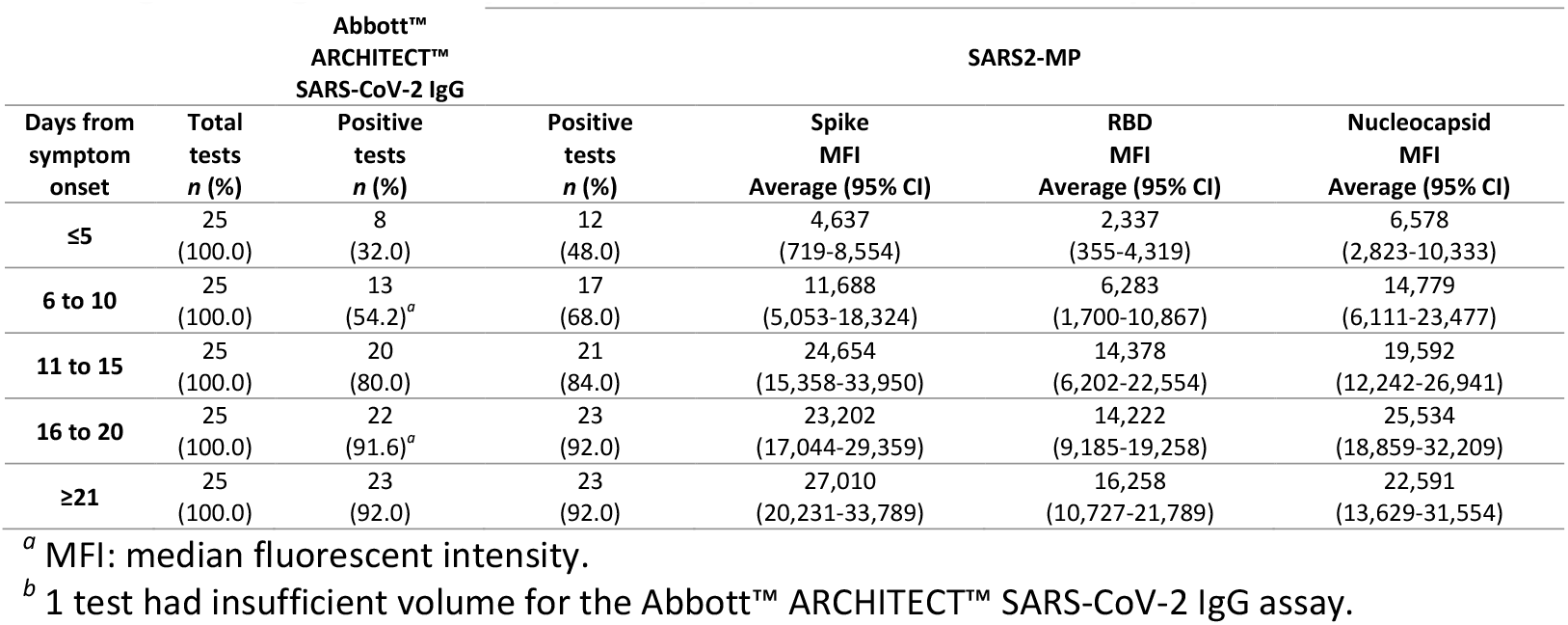
Percent positivity and average MFI^*a*^ values of 3Flex *vs*. Abbott™ ARCHITECT™ SARS-CoV-2 IgG serological tests by days from symptom onset for 125 unique patients.

### Serological profiling of SARS-CoV-2 infection

#### Characteristics of patient population

All 140 unique patients profiled were positive for SARS-CoV-2 nucleic acid by RT-PCR. Of these, 65/140 (46.4%) were women and 75/140 (53.6%) were men **(Table 3)**. Sixty-six patients were admitted to an ICU, and there were 20 mortalities—with 16/20 (80%) being men. Nursing home residents comprised 37/140 (26.4%) of the tested patients, and the average age of nursing home residents was 82.6 and 76.0 years for women and men, respectively, compared to 63.4 and 64.1 years for other patient types (inpatient, outpatient) combined. The most frequently reported symptoms were fever (women: 32/65, 49.2%; men: 51/75, 68.0%) and shortness of breath (women: 34/65, 52.3%; men: 44/75; 58.7%). Gastrointestinal symptoms (diarrhea/vomiting) were noted for 42/140 (30.0%) of patients (women: 16/65, 24.6%; men: 26/75, 34.7%). Ventilated patients comprised 47/140 (33.6%) of the population (women: 22/65, 33.8%; men: 25/75, 33.3%). The most frequently reported comorbidities included hypertension (women: 45/65, 69.2%; men: 55/75, 73.3%), a history of smoking (women: 33/65, 50.8%; men: 44/75, 58.7%), and diabetes (women: 23/65, 35.4%; men: 33/75, 44.0%).

**Table 3:** Demographics, symptoms, and co-morbidities of 140 patients positive for SARS-CoV-2 by RT-PCR and tested for IgG in this study.

| **Demographics** | **n** | **(%)** |
| --- | --- | --- |
| Female | 65 | (46.4) |
| Male | 75 | (53.6) |
| Nursing home residents | 37 | (26.4) |
| Age (average) in years | 68.0 |  |
| **Symptoms** |  |  |
| Fever | 83 | (59.3) |
| Cough | 64 | (45.7) |
| Headache | 12 | (8.6) |
| Sore throat | 10 | (7.1) |
| Shortness of breath | 78 | (55.7) |
| Altered mental status | 45 | (32.1) |
| Malaise | 36 | (25.7) |
| Diarrhea / Vomiting | 42 | (30.0) |
| **Course of disease / Complications** |  |  |
| Days of hospitalization (average) | 13.8 |  |
| ICU admission | 66 | (47.1) |
| Low oxygen saturation | 75 | (53.6) |
| Ventilation | 47 | (33.6) |
| Septic shock / vasopressor | 28 | (20.0) |
| Bacterial pneumonia | 28 | (20.0) |
| Deceased | 20 | (14.3) |
| **Comorbidities** |  |  |
| Asthma | 15 | (10.7) |
| COPD | 13 | (9.3) |
| Diabetes | 56 | (40.0) |
| Hypertension | 100 | (71.4) |
| Obstructive sleep apnea | 24 | (17.1) |
| Coronary artery disease | 19 | (13.6) |
| Cerebrovascular disease | 25 | (17.9) |
| Immunodeficiency / Immunosuppression | 11 | (7.9) |
| History of smoking | 77 | (55.0) |
| **Total** | **140** | **(100)** |

#### *Serological response to* SARS-CoV-2 *S, RBD, and NP antigens*

MFI values corresponding to each antigen were measured for all sera tested (*n* = 534) and analyzed with respect to days from symptom onset **(Figure 1B, Table 4)**. The peak IgG response to S and RBD antigens occurred on 23.8 days and 23.6 days from symptom onset, respectively. The peak IgG response to NP was observed at 16.7 days from symptom onset. No test was qualitatively interpreted as negative >28 days from symptom onset. Although MFIs for all antigens decreased following the peak response, all seroconverted patients with at least one test following a positive (*n* = 59) were interpreted as positive on subsequent tests with the exception of three patients with discrepant findings. Two of these three patients were mortalities with only limited sera available (i.e. two samples) from a narrow timeframe (<11 days from symptom onset). The serological response of the third discrepant patient (Patient H) was profiled later **(Figure 2)**.

**Table 4:** Percent positivity and average MFI^*a*^ values of all 3Flex serological tests by days from symptom onset for all patients tested^*b*^

| Days from symptom onset | Total tests<br>*n* (%) | Negative tests<br>*n* (%) | Positive tests<br>*n* (%) | Spike MFI<br>Average (95% CI) | RBD MFI<br>Average (95% CI) | Nucleocapsid MFI<br>Average (95% CI) |
| --- | --- | --- | --- | --- | --- | --- |
| ≤5 | 67<br>(100.0) | 33<br>(49.3) | 34<br>(50.7) | 11,565<br>(5,718-17,412) | 8,723<br>(3,295-14,150) | 13,794<br>(8,721-18,866) |
| 6 to 10 | 106<br>(100.0) | 21<br>(19.3) | 85<br>(80.2) | 23,051<br>(18,955-27,147) | 16,172<br>(12,323-20,023) | 27,028<br>(21,749-32,308) |
| 11 to 15 | 85<br>(100.0) | 9<br>(10.6) | 76<br>(89.4) | 29,545<br>(25,190-33,901) | 19,876<br>(15,534-24,217) | 28,598<br>(23,978-33,218) |
| 16 to 20 | 88<br>(100.0) | 5<br>(5.7) | 83<br>(94.3) | 31,006<br>(26,406-35,606) | 20,694<br>(16,539-24,848) | 28,474<br>(24,134-32,814) |
| ≥21 | 188<br>(100.0) | 5<br>(2.7) | 183<br>(97.3) | 31,751<br>(28,774-34,727) | 19,516<br>(16,889-22,142) | 20,459<br>(17,574-23,343) |

<sup>a</sup> MFI: median fluorescent intensity.
<sup>b</sup> Includes multiple serological tests from non-unique patients.

**Figure 1.**
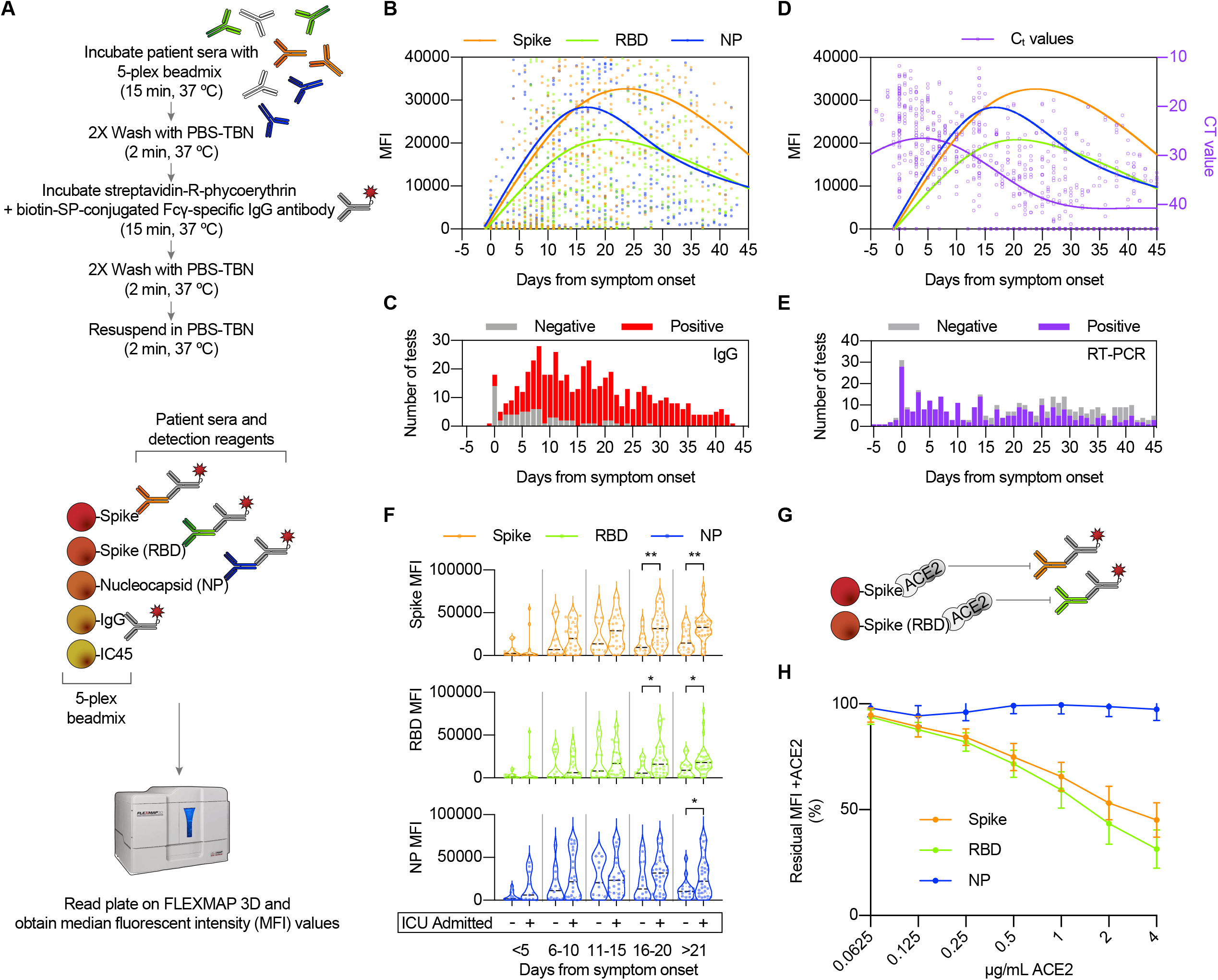
Utility of the 3Flex multiplex microsphere-based immunological assay for SARS-CoV-2 diagnostics and immunological research. **(A)** Simplified workflow and illustration of the 3Flex assay. **(B)** Total composite profile of IgG immune reactivity in SARS-CoV-2 -infected patients by days from symptom. Data represented are median MFI values for each viral antigen (Spike, RBD, and NP) from 534 serological tests (1,602 data points) of 140 patients. Tests shown are those occurring between −5 to 45 days from symptom onset and includes tests deemed qualitatively negative or positive. Data points outside the axis limits (273) are not shown. Fitted curve lines are smoothed splines with 4 knots. **(C)** Histogram showing number of serological tests by day from symptom onset and their qualitative positivity. **(D)** Composite profile of cycle threshold (C_t_) values from the same SARS-CoV-2 infected patients overlaid with the serological profile. C_t_ values are plotted in reverse on the right axis (purple). A total of 521 RT-PCR tests are depicted and each has 2 data points plotted. Undetected targets or negative tests were assigned a C_t_ of 45. Fitted curve lines are smoothed splines with 4 knots. **(E)** Histogram showing number of RT-PCR (molecular) tests by day from symptom onset and their qualitative positivity. **(F)** Violin plot comparison of median MFI values for patients admitted to the ICU (*n* = 66) *vs*. all other SARS-CoV-2-infected patients (*n* = 74). Data shown are for serological tests interpreted as positive only. Only 1 test for each unique patient was randomly selected for inclusion in each time interval bin (≤5, 6 to 10, 11 to 15, 16 to 20, and ≥21 days from symptom onset). Differences between violin plots are calculated using an unpaired *t* test: *, P ≤ 0.05; **, P ≤ 0.01. **(G)** Assay schematic for ACE2 inhibition of antibody binding to the SARS-CoV-2 S and RBD antigens. **(H)** Addition of ACE2 inhibits detection and binding of antibodies to S and RBD, but not to NP. Percent residual (i.e. compared to no ACE2 added) mean MFI values with standard deviation are shown at increasing ACE2 concentrations (*x*-axis log2 scale). Sera tested (*n* = 11) were from patients positive for SARS-CoV-2 IgG.

**Figure 2.**
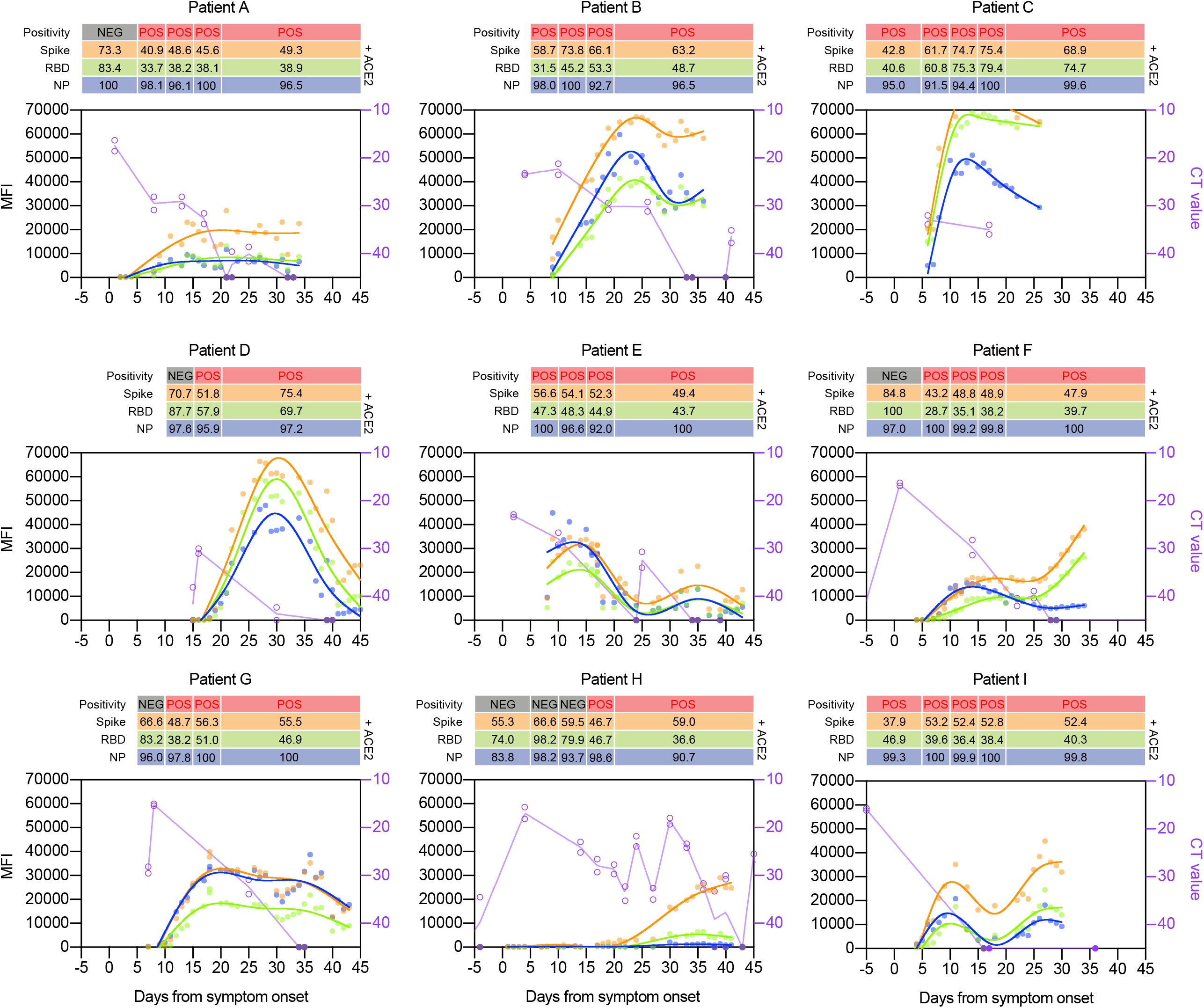
Interperson variation in longitudinal IgG immunological responses to SARS-CoV-2 in intensively hospitalized patients. Longitudinal serum samples from ICU-admitted patients (*n* = 9) were tested for IgG to SARS-CoV-2 S, RBD, and NP antigens. Curve lines were fitted for each antigen (smoothed splines with 4 knots). Corresponding RT-PCR C_t_ values were plotted in reverse on the right axis (purple) and connected by straight lines through the mean C_t_ value of each test. A single sample was randomly selected for each time interval bin (≤5, 6 to 10, 11 to 15, 16 to 20, and ≥21 days from symptom onset) to assay for ACE2 inhibition. Values shown above each plot are residual MFI values (in percent) for each antigen (percentage of MFI detected with ACE2 addition compared to a no-ACE2 control).

Overall, 61/534 tests were interpreted as negative and 473/534 tests were interpreted as positive **(Figure 1C, Data S1)**. Twelve of 534 tests were interpreted as positive based on one antigen MFI value exceeding the threshold for positivity (S: 9/12, 75%; RBD: 3/12, 25%; NP: 0/12, 0.0%). For 57 tests interpreted as positive based on two antigens, the frequency of antigen pairs were S and RBD (52/57, 91.2%), S and NP (4/57, 7.0%), and RBD and NP (1/57, 1.8%). Single- and double-antigen positives occurred at a median of 5.5 and 8.0 days from symptom onset, respectively. When allowing for only one test (*n* = 231) per unique patient (*n* = 140) in each time interval the IgG positivity rate was 53.4% when serum was collected ≤5 days from symptom onset, and 96.1% when collected ≥21 days from symptom onset (**Table 3)**.

C_t_ values (*n* = 1,042; 2 C_t_ values per RT-PCR test) were obtained from RT-PCR detection of SARS-CoV-2 nucleic acid and compared to the serological response to S, RBD, and NP **(Figure 1D)**. Nucleic acid detection peaked (i.e. lowest average C_t_ values) at 4.6 days from symptom onset. For tests conducted <21 days from symptom onset 85.2% were positive and had a mean C_t_ value of 26.3 ± 0.4 (SEM) **(Figure 1E)**. In contrast, 47.8% tests conducted ≥21 days from symptom onset were positive and had a mean C_t_ value of 33.4 ± 0.4 (SEM).

#### Serological response to SARS-CoV-2 in ICU-admitted patients

Patient populations were assessed for differences in MFI values obtained for each SARS-CoV-2 antigen. Higher MFI values were observed for patients aged <65 (*vs*. ≥65, not shown), and for those admitted to an ICU **(Figure 1F)**. ICU-admitted patients demonstrated significantly higher MFI values for all SARS-CoV-2 antigens at >21 days from symptom onset. ICU-admitted patients (*vs*. not-admitted patients) were not appreciably different with respect to age (67.0 years *vs*. 68.9 years), sex (49.5% female *vs*. 46.9% female), or histories of smoking, cerebrovascular disease, coronary artery disease, hypertension, diabetes, or asthma. Patients with histories of chronic obstructive pulmonary disease (10/66 *vs*. 3/74, P = 0.0383, Fisher’s exact test) and obstructive sleep apnea (16/66 *vs*. 8/74, P = 0.0438) were more frequent in the ICU-admitted patient population than in non-admitted individuals.

#### Inhibition of SARS-CoV-2 antibody binding to S and RBD using ACE2

To assess whether 3Flex could assay for antibodies that neutralize receptor binding, titrated concentrations (0.0625 – 4 μg/mL) of ACE2 were incubated with the 5-plex beadmix prior to addition of serum **(Figure 1G)**. The addition of ACE2 resulted in loss of detection of S and RBD antibodies, but not NP **(Figure 1H)**. The 2 μg/mL concentration of ACE2 was selected for the following studies because it was capable of producing a >50% drop in anti-RBD IgG detection. The residual MFI detected at 2 μg/mL of ACE2 was 53.1% ± 8.0%, 43.5% ± 9.72%, and 98.6% ± 4.7%, for S, RBD, and NP, respectively (mean ± SD).

#### Longitudinal serological response to SARS-CoV-2 in extensively sampled individuals

Nine patients (6 women, 3 men) were extensively sampled due to extended hospitalization (all admitted to an ICU) for COVID-19. The IgG response to SARS-CoV-2 was explored in each of these, alongside ACE2 inhibition testing **(Figure 2)**. The average age of these patients was 67.3 years (range: 47.0 to 80.0 years). All of these patients survived their illness. All patients were ventilated except for Patient I. There was a wide-ranging prevalence of comorbidities and COVID-19 complications present in this group. These patients had histories notable for hypertension (9/9), diabetes (7/9), smoking (5/9), kidney disease (7/9). Where available, C_t_ values typically decreased or were undetectable following a peak IgG response. Highly variable (MFI magnitude and kinetics) serological responses were observed for these individuals. In addition, ACE2 inhibition was demonstrated to varying degrees for each patient, generally with the greatest ‘neutralization’ observed for sera tested ≥21 days from symptom onset. Neutralization was exclusive to S and RBD, with NP showing no significant inhibition. Despite evidence of an IgG response, and having tested negative by RT-PCR, 4/9 patients subsequently tested RT-PCR positive for SARS-CoV-2 on later follow-up testing.

## DISCUSSION

In this work we have validated 3Flex as a multiplex fluorescent microsphere immunoassay to qualitatively assess the IgG response to infection by SARS-CoV-2. We found that this assay compared favorably with the Abbott™ ARCHITECT™ SARS-CoV-2 IgG assay when testing serum from a diverse group of individuals with a spectrum of COVID-19 clinical presentations. The overall profile of the IgG serological response in this collection revealed the initial and peak responses for each SARS-CoV-2 antigen, timing of the decline in antibody levels, and correlation with decline in Ct values (a proxy for viral load).

Through the early stages of the pandemic, the kinetics of the antibody response to SARS-CoV-2 were described using a variety of methodologies and antigenic targets. Although it is challenging to directly compare results, the overall picture which emerges from the literature is that most individuals develop IgM, IgA, and IgG responses at approximately 5-14 days post symptom onset, though some can take up to 14-28 days ^22-25^. In this study, we showed similar kinetics. Across 534 tests from 140 individuals 48% of those with reported symptom onset within the past 5 days had detectable IgG, but that number rose to 68% by days 6-10 and 92% by days 16 to 20. Further, as the pandemic enters its ninth month since the first cases were identified in the US, some studies have raised concerns over declining levels of SARS-CoV-2 antibodies and the potential ramifications for natural immunity and vaccine response ^26^. Among the nine patients for who we had longitudinal samples, none of the individuals became negative according to the qualitative interpretation of the assay. In addition, several individuals who were tested at later time points (>50 days post symptom onset) also demonstrated lower but still qualitatively positive results (data not shown). These findings are consistent with serological data from Iceland which indicated that infected individuals remained seropositive up to 4 months post-infection ^27^.

Whereas many serological assays for COVID-19 detect a single antigen, this assay incorporates S, RBD, and NP. This feature may prove useful as it has the added flexibility to potentially discriminate between the immune responses of infected or vaccinated people (albeit, a use entirely dependent on the ultimate composition of upcoming SARS-CoV-2 vaccines). This multi-target approach may also improve the sensitivity of the assay, since a positive result for any of the antigens is interpreted as positive overall. We observed that ∼15% of samples were positive for one antigen but not another, though after 28 days from symptom onset the majority of patients were positive for all antigens. Here, detection of S and RBD increased the sensitivity of the 3Flex assay among some of the earlier samples (≤5 days from symptom onset) when compared to the NP-only Abbott™ ARCHITECT™ SARS-CoV-2 IgG assay. Currently, there are mixed reports concerning antigen specific dynamics of the antibody response, i.e. the relative timing and duration of responses to one viral antigen versus another. Despite similar findings reported by Liu et. al. ^28^, others have reported greater sensitivity at earlier time points using single-target NP-based assays.^29^

The kinetics of the antibody response were not appreciably different by demographic measures such as age or sex. Higher MFI values were observed in patients admitted to the ICU as part of their COVID-19 disease course. This association of higher antibody levels with disease severity has been reported by others ^30,31^, though our data suggested that antigen-specific antibody kinetics did not differ significantly between the two groups—i.e. in aggregate, peak responses were observed at the same day from symptom onset.

A limitation of this study is its dependence on retrospective and remnant serum samples which hindered longitudinal sampling. However, for a subset of nine individuals admitted to the ICU, we had multiple samples that enabled us to track their antibody and viral load data over time. This allowed us to look beyond the composite data **(in Figure 1B)** for a depiction of individual responses **(Figure 2)**. The longitudinal MFI values of these individual patients suggested a high degree of interperson variation in terms of both kinetics and relative antigen responses. For most of these nine patients the MFIs for anti-S antibodies were higher. This is despite the fact that S protein was coupled to beads at only 10% of the molarity of RBD or NP. Thus, anti-S antibodies potentially comprise a larger proportion of anti-SARS-CoV-2 antibodies. We observed interperson and time (from symptom onset)-dependent variability in the ability of soluble ACE2 to block the binding of anti-S and anti-RBD antibodies, an indirect measure of neutralizing capability. In general, a trend towards greater neutralization over time from symptom onset was observed. The ability to measure inhibition of signal with ACE2 may prove useful to measure the effectiveness of, and discriminate between, the immune responses of infected or vaccinated individuals. However, an important caveat is that it is unclear how *in vitro* ‘neutralization’ performed in this manner translates to *in vivo* immunity.

In summary, the 3Flex assay described here is a versatile and robust method to measure antibodies to the major antigens of SARS-CoV-2. Beyond the current pandemic, the xMAP microsphere technology allows for rapid and flexible assay design, as well as straightforward implementation. This provided a serological platform capable of responding to new and emerging pathogens.

## Data Availability

All manuscript data is available as an attached data set.

## ACKNOWLEDGMENTS

We thank the UR Medicine Central Laboratory Clinical Microbiology for specimen collection and serological testing.

## CONTRIBUTIONS

AC and NP designed the study, selected sera, and performed statistical analyses. SA provided technical expertise with assay design, reagent selection, and assay optimization. SA, AC, and NP interpreted criteria for assay validation. MC provided sera from confirmed respiratory disease infections for interference testing. JW and MZ provided technical expertise and additional reagents. AC, CP, JB, ABC, and NP reviewed patient charts for demographic information and COVID-19 histories. LB, ZP, AC, NP, and SA performed serological testing. DH and NP provided laboratory resources. AC and NP analyzed data/results and wrote the first draft of the manuscript. All authors participated in editing and reviewing the manuscript and approved the final manuscript. All authors contributed to the article and approved the submitted version.

